# Healthy Dietary Patterns and Lower Atherosclerosis in High-Risk Individuals

**DOI:** 10.1101/2024.11.19.24317591

**Authors:** Laurel Cherian, Puja Agarwal, Sonal Agrawal, Bryan D. James, Dixon Yang, Maude Wagner, Sue Leurgans, David Bennett, Neelum Aggarwal, Julie Schneider

## Abstract

**Background:** This study investigates associations between diet patterns and the risk of intracranial atherosclerosis (ICAD) in individuals with preexisting hypertension (HTN) or myocardial infarction (MI).

**Method:** 676 autopsied participants (mean age at death =91.1±6.1, 71% women) of a longitudinal clinical neuropathological cohort study, with complete dietary and neuropathology data, were included. Diet scores were computed (median interval to death = 5.9 (3.0, 8.7 years). HTN and MI history was self-reported. Large vessel atherosclerosis was evaluated at the circle of Willis, and severity of intracranial atherosclerosis was assessed based on number of atherosclerotic plaques, extent of vessel involvement, and degree of vessel occlusion to create a 4-level grading system (0-3). All regression models were adjusted for age, sex, education, caloric intake, and APOE4.

**Results:** Of the 676 subjects, 361 (53%) had mild, 142 (21%) had moderate, and 29 (5%) had severe atherosclerosis. There was no direct relationship between diet and atherosclerosis. The relationship between ICAD and MI (OR = 1.38, 95% CI = 0.95, 2.00) showed a nonsignificant trend. HTN (OR = 1.598, 95% CI = 1.15, 2.18) was positively associated with intracranial atherosclerosis. The association of diet with intracranial atherosclerosis differed by history of MI (MIND (p=0.007), MedDiet (p=0.006)). The association between ICAD and the MIND diet also differed by whether HTN was reported (β=-0.212, SE= 0.111, p=0.055) as did the relationship between ICAD and the MedDiet (β = -0.077, SE= 0.035, p=0.029). In stratified analysis, among individuals with preexisting MI (N=130), those with a better diet had lower odds of intracranial atherosclerosis (MedDiet: OR = 0.88, 95% CI = 0.81, 0.96; MIND: OR = 0.69, 95% CI = 0.53, 0.90).

**Conclusion:** A healthy dietary pattern is associated with lower odds of severe intracranial large vessel atherosclerosis in high-risk older adults. In-vivo studies of dietary habits and brain health, specifically in those at high vascular risk are needed.

## INTRODUCTION

Intracranial atherosclerotic disease (ICAD) is prevalent in community-based cohorts.^1^ It is well established that ICAD causes ischemic stroke, with large-artery intracranial atherosclerosis implicated as the etiology in up to 30% of all ischemic stroke cases, and even more frequently in African and Asian populations.^2^ Ischemic strokes have the highest recurrence rate of all stroke subtypes, with a 10%–24% annual risk of stroke recurrence, despite optimized medical treatment.^3, 4^ In addition to its sizable contribution to the global burden of stroke, intracranial atherosclerosis has been associated with faster cognitive decline and increased risk of dementia, even in stroke-free individuals.^5, 6^ As such, identifying key modifiable risk factors for intracranial atherosclerosis, preventing its development, and implementing interventions to slow its progression is important.

Although prior epidemiological studies have shown that ICAD is more common in individuals with uncontrolled vascular risk factors,^7, 8^ there are relatively little data on the pathophysiology of ICAD compared to extracranial carotid artery atherosclerosis or coronary artery atherosclerosis. One modifiable risk factor that may impact ICAD is diet. The Mediterranean-DASH intervention for neurodegenerative delay (MIND) diet, which combines the Mediterranean diet and Dietary Approaches to Stop Hypertension diet, and targets foods associated with cognitive function in later life, has been found to improve cardiovascular health, slow cognitive decline and reduce dementia risk.^1, 9, 10^ In a recent observational cohort study, the MIND diet was associated with fewer cardiovascular events, including stroke, assessed over a 10 year of follow-up period.^11^ Further, a recent case-control study found the highest alignment with the MIND diet associated with a 59% reduction in stroke risk.^12^

Other studies have found similar results regarding the benefits of a healthy diet in reducing cardiovascular disease in individuals with elevated vascular risk. In the Lyon Diet Heart Study, survivors of myocardial infarction currently adhering to a Mediterranean diet (MedDiet) experienced a 70% reduction in stroke and recurrent myocardial infarction after four years, compared to those following a “prudent Western” diet.^13^ This was more than twice the effect of simvastatin in the Scandinavian Simvastatin Survival study, which also studied survivors of myocardial infarction.^14^ The MedDiet was also found to be protective in preventing cardiovascular events, including stroke, in high-risk individuals in the PREDIMED study.^15^

Although the associations among diet, vascular risk factors, and cardiovascular events are well-established, the role of diet in the development of ICAD in older brains is less clear, and even less so in individuals with pre-existing vascular risk factors, such as hypertension or a history of myocardial infarction. Most studies on ICAD have utilized neuroimaging, with fewer studies examining neuropathology.^16^ This study investigates the associations between diet patterns (MIND diet and MedDiet) and ICAD in older at-risk individuals with pre-existing vascular risk factors, specifically history of hypertension and myocardial infarction. To this end, we leveraged dietary assessments and postmortem neuropathology from older, deceased participants with autopsies in the Rush Memory and Aging Project (MAP) to examine the associations between diet and ICAD in high-risk individuals.

## METHODS

### Study Participants

The Rush MAP is an ongoing, longitudinal, clinical–pathologic cohort study of aging and dementia, which started in 1997 and enrolls women and men from more than 40 retirement communities and housing facilities in the metropolitan Chicago area.^17^ Eligibility requires the absence of known dementia and agreement to annual clinical evaluations and brain donation at death. To date, the follow-up rate exceeds 90%, and the autopsy rate exceeds 80%. All participants signed an informed consent and an Anatomical Gift Act for organ donation and the study protocol was approved by an Institutional Review Board of Rush University Medical Center.

### Eligible participants for analysis

Of the 2,198 MAP participants who had completed the baseline clinical evaluation at the time of these analyses, we first identified 2,042 individuals alive and active when the dietary sub study began in 2004. We then excluded 162 participants without FFQ data during the follow-up (9 had withdrawn, 83 died, and 70 were Spanish speakers, had a clinical diagnosis of dementia, or declined to enroll in the dietary sub study), 486 with FFQ data unprocessed or incomplete, and 4 with FFQ data under quality check. Lastly, we excluded 577 participants still alive, 111 decedents without complete brain autopsy to date, and 26 decedents who did not yet have data on intracranial vascular neuropathology, and one participant missing HTN and MI history, leaving an analytic sample of 676 participants.

### Dietary Assessment

In MAP, dietary intake assessments began in February 2004, and has continued annually thereafter using a validated >144-item semiquantitative FFQ, modified from the Willett for use older adults; validity and reliability of this FFQ has been established.^18^ For each food item, participants were asked to report the usual frequency of intake during the past year. For each food item in the FFQ, total calories and nutrient levels were based either on natural portion sizes (e.g., one apple) or according to age- and sex-specific portion sizes from national dietary surveys, as previously defined.^9^ In this study, we examined average MIND and MedDiet scores calculated using all available repeated FFQs assessed until death.

### MIND Diet Score

The MIND diet score is comprised of 15 dietary components, each of which is summed to produce a score ranging from 0-15, with a higher score reflecting higher concordance.^9^ 10 brain-healthy food groups (green leafy vegetables, other vegetables, nuts, berries, beans, whole grains, fish, poultry, olive oil, and wine) and 5 unhealthy food groups (red meats, butter and stick margarine, cheese, pastries and sweets, and fried foods). Consumption of the recommended amount or greater of the healthy food items were scored as a “1,” while consumption of unhealthy food items was reverse coded, so that the “1” was scored if the participant consumed less of the item and “0” if they consumed more.^9^

### Mediterranean Diet Score

The Mediterranean diet score, as described by Panagiotakos et al.,^19^ was also computed. This score is comprised of 11 dietary components, using serving sizes based on the traditional Greek Mediterranean diet as the comparison metric. Each component is scored 0 to 5, and all are summed for a total score ranging from 0 to 55 (highest dietary concordance).^10^

### Intracranial Atherosclerosis Disease Outcome

Methods for brain autopsy and pathologic evaluation for vascular pathology have been previously described.^20^ Briefly, ICAD assessment was completed by visual inspection of the circle of Willis, including the basilar, posterior cerebral, middle cerebral, and anterior cerebral arteries and their proximal branches. Vessels were visually inspected to determine the number of plaques, extent of involvement of each artery by plaques, and the degree of stenosis on bisecting vessels.^21^ A 4 level scale was created for analysis with 0 for no significant ICAD observed; 1 for mild atherosclerosis with small amounts in up to several arteries (typically less than 25% vessel involvement) without significant occlusion; 2 for moderate atherosclerosis in up to half of all visualized major arteries, with less than 50% occlusion of any single vessel; and 3 for severe atherosclerosis in more than half of all visualized arteries, and/or more than 75% occlusion of one or more vessels. In this study, we considered the presence of ICAD (none-mild vs. moderate-severe) based on histologic examination using hematoxylin & eosin–stained sections of the anterior basal ganglia.^21^

### Hypertension and myocardial infarction

History of hypertension (HTN) was defined based on self-report and medication review.^22^ History of myocardial infarction (MI) was determined by the physician on the basis of a uniform structured neurological examination and clinical history.

### Other covariates

Covariates examined included sociodemographic (age at death, sex, years of formal education) and the presence of apolipoprotein E ɛ4 (APOE4) genotyping (at least 1 allele).^23^ Smoking (never smoked, former smoker, or current smoker), and years of formal education, which were reported at enrollment. History of diabetes was defined based on self-report and medication review.^22^ History of stroke and heart failure were determined by the physician on the basis of a uniform structured neurological examination and clinical history.

### Statistical analysis

Spearman correlation was used to examine the correlation of MedDiet and MIND diet. Next, for each dietary pattern, we used a logistic regression model to test the hypothesis that greater dietary scores, reflecting healthier dietary habits, were associated with lower odds of ICAD at death, after controlling for age at death (continuous, years), sex, education (continuous, years), calories intake (continuous, kcal/day), and APOE4 status. In addition, we also used separate logistic regression models to test the hypothesis that history of HTN and MI were associated with ICAD, adjusted for age at death, sex and education.

Next, to examine the interaction between continuous diet scores and history of MI, we fitted separate logistic regression models with ICAD as the outcome and terms for the continuous diet score, history of HTN, and a multiplicative term of these two in our model adjusted for age at death, sex, education, calories intake, and APOE4 status. Similar analyses were repeated for history of HTN. For Figure 1, we assessed the MIND diet and MedDiet association with mild, moderate and severe ICAD using natural cubic splines among those with and without history of heart disease

**Figure 1:**
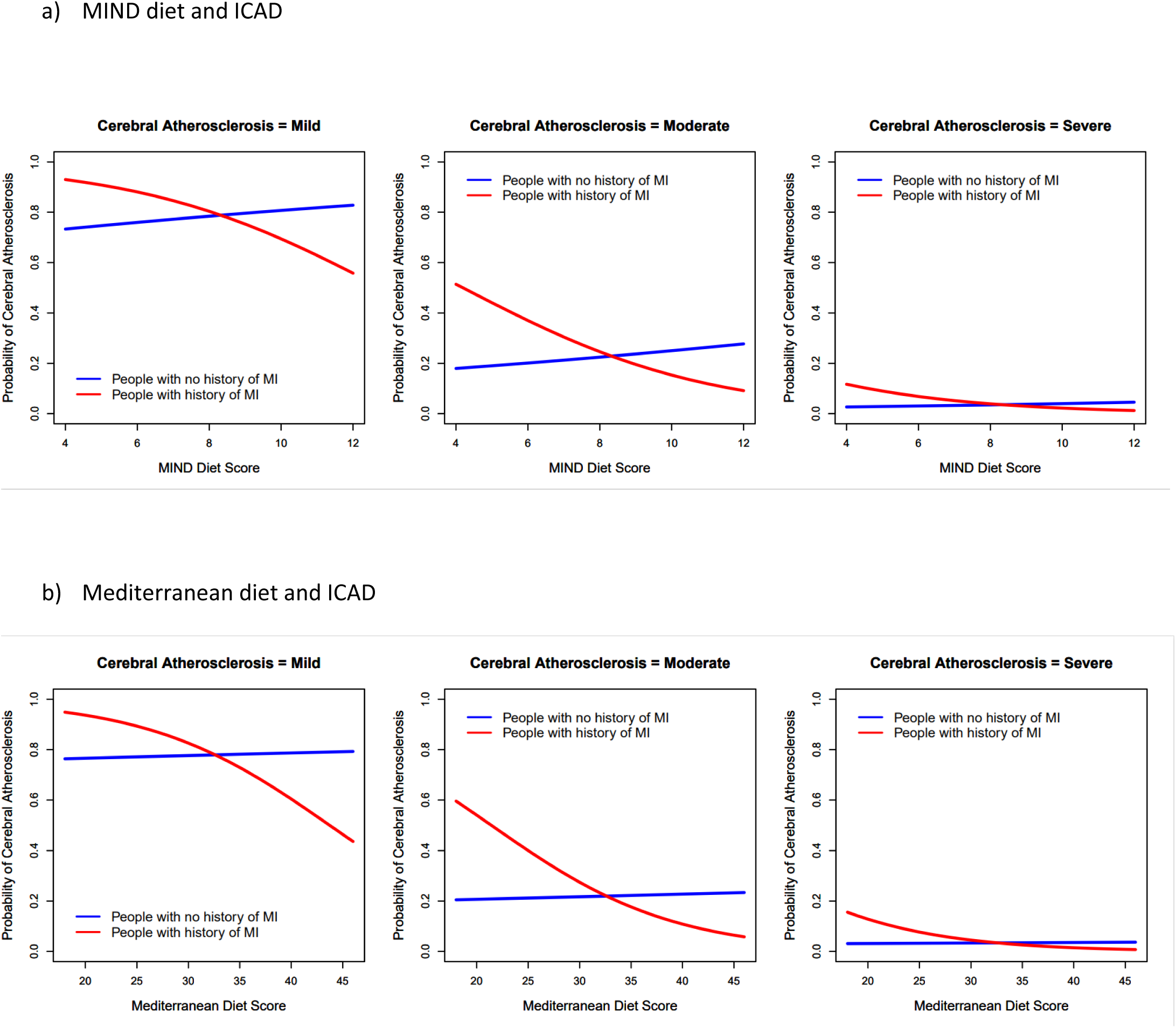
Association of a) MIND and b) MedDiet with mild, moderate, and severe intracranial atherosclerotic disease (ICAD) among those with and without a history of heart disease.

Lastly, to provide a clinical context and a public health inference, we further dichotomized the cohort using the median diet score (7.5 for the MIND diet and 30 for the MedDiet) into a low diet score group, representing those with a less healthy diet and a high diet score group, representing those with a healthier diet. This was done for both MIND and MedDiet scores. Using logistic regression models, we assessed the interaction of low/high diet and vascular risk (HTN and MI) for the association of these vascular risk factors with ICAD. When interactions were significant, we ran models for the association of diet with ICAD stratified by HTN/MI.

Sensitivity analyses were performed in models with the interaction terms by additional adjustment for potential confounders to assess the robustness of significant outcomes. We further adjusted for clinical vascular risk factors, including diabetes, stroke, heart failure, smoking, and then ICAD, one at a time. All statistical analyses were conducted using SAS software version 9.4 (SAS Institute, Cary, NC).

## RESULTS

The analytic sample included 676 MAP participants with a mean age at death of 91.1 (SD=6.1) years, and this was similar across the HTN and MI groups analyzed (Table 1). 197 (29%) of subjects were male, with higher percentages of males in the group without a history of HTN (39%) and in those with a history of MI (42%). Participants averaged 14.8 years (SD=2.8) of education, independent of vascular risk factors. Mean MIND and MedDiet scores, assessed over a mean follow-up of 5.85 (SD=3.92) years before death, were 7.4 (SD=1.4) and 30.1 (SD=4.5), respectively, and were highly correlated (rho= 0.69, p<.0001). Of the 676 selected subjects, 361 (53%) had mild, 142 (21%) had moderate, and 29 (5%) had severe ICAD.

**Table 1.** Demographic, clinical, and neuropathologic characteristics by presence or absence of hypertension (HTN) and myocardial infarction (MI). Values are expressed as mean (SD), unless otherwise specified.

| Demographic, clinical, and neuropathologic characteristics | Overall (N=676) | No HTN (n=210) | History of HTN (n=466) | No MI (n=544) | History of MI (n=132) |
| --- | --- | --- | --- | --- | --- |
| Demographics |  |  |  |  |  |
| Age at death, years | 91.1 (6.1) | 91.7 (6.0) | 91.0 (6.2) | 91.2 (6.2) | 91.0 (5.9) |
| Male, no. (%) | 197 (29) | 81 (39) | 115 (25) | 141 (26) | 55 (42) |
| Education, years | 14.8 (2.87) | 14.8 (3.1) | 14.9 (2.8) | 14.8 (2.8) | 15.1 (3.0) |
| APOE4 carriers (at least 1 allele), no. (%) | 142 (21) | 47 (22) | 95 (20) | 123 (23) | 19 (14) |
| Diet* |  |  |  |  |  |
| MIND Diet score (possible score 0-15) | 7.4 (1.4) | 7.4 (1.4) | 7.4 (1.4) | 7.4 (1.4) | 7.5 (1.5) |
| MedDiet score (possible score 0-50) | 30.1 (4.5) | 30.8 (4.6) | 29.8 (4.5) | 30.2 (4.6) | 29.9 (4.2) |
| Total calories, kcal/day | 1843.6 (528.5) | 1894.7 (547.3) | 1832.0 (524.0) | 1813.6 (515.6) | 1912.1 (552.2) |
| Vascular risk factors<sup>†</sup>, no. (%) |  |  |  |  |  |
| History of MI | 132 (19) | 38 (18) | 94 (20) | - | - |
| History of HTN | 466 (69) | - | - | 372 (68) | 94 (71) |
| History of diabetes | 86 (13) | 11 (5) | 75 (16) | 372 (68) | 94 (71) |
| History of stroke | 133 (20) | 32 (15) | 101 (22) | 64 (12) | 22 (17) |
| History of congestive heart failure | 94 (14) | 20 (10) | 74 (16) | 58 (11) | 36 (28) |
| Intracranial atherosclerosis disease, no. (%) |  |  |  |  |  |
| Absent | 144 (21.27) | 59 (28.10) | 85 (18.24) | 124 (22.79) | 20 (15.15) |
| Mild | 361 (53.32) | 107 (50.95) | 254 (54.51) | 290 (53.31) | 71 (53.79) |
| Moderate | 142 (20.97) | 33 (15.71) | 109 (23.39) | 107 (19.67) | 35 (26.52) |
| Severe | 29 (4.28) | 11 (5.24) | 17 (3.65) | 23 (4.23) | 5 (3.79) |

Abbreviations: APOE4, Apolipoprotein E epsilon 4; BP, blood pressure; MIND, Mediterranean-DASH Intervention for Neurodegenerative Delay; MedDiet, Mediterranean Diet. One person has missing data on the history of hypertension and MI.
\* Dietary scores and total calories represent mean values calculated over the entire follow-up until death.
<sup>†</sup> Vascular risk factors assessed at baseline and throughout follow-up.

**Table 2:** Association of diet scores, HTN, Myocardial infarction with ICAD.

| Model | Model term | Odd Ratio (95%CI) | P Value |
| --- | --- | --- | --- |
| **Overall Sample** |  |  |  |
| **A**<sup>a</sup> | MIND score | 1.01 (0.90, 1.11) | 0.966 |
| **B**<sup>a</sup> | MedDiet score | 0.98 (0.94;1.01) | 0.235 |
| **C**<sup>b</sup> | History of MI | 1.39 (0.96, 2.01) | 0.079 |
| **D**<sup>b</sup> | History of HTN | 1.55 (1.13, 2.13) | 0.006 |

<sup>a</sup>Models were adjusted for age, sex, education, calories, and APOE4. <sup>b</sup>Models were adjusted for age, sex, education.

### Association of Diet, HTN, and MI with ICAD

Controlling for demographics, caloric intake, and APOE4 status, we did not find evidence that greater MIND diet scores (Odd Ratio [OR]=1.01; 95% CI = 0.90,1.11; p=.966) and greater MedDiet scores (OR=0.98; 95%CI=0.94,1.01; p=.235) were associated with lower odds of ICAD (Table 1). In addition, controlling for demographics, history of MI was not associated with ICAD (OR=1.39; 95%CI= 0.96, 2.01; p=.079) (Table 1). However, history of HTN was associated with 55% higher odds of having intracranial atherosclerosis at death (OR=1.55; 95%CI=1.13, 2.13; p=.006), after controlling for demographics (Table 1).

### Interaction Between diet and vascular risk factors for the severity of intracranial atherosclerosis

To examine whether the history of vascular risk factors modified the association between MIND diet and atherosclerosis, regression models included an interaction term between MIND diet and the presence of history of MI or HTN in separate models (Table 3). The interaction term for MIND*MI was significant for ICAD (OR=0.69; 95%CI=0.53, 0.91); p = 0.007), indicating that MIND diet was associated with reduced risk of atherosclerosis in people with history of MI than for people without history of MI. Similarly, Med Diet*MI interaction term was also significant (Table 3).

**Table 3:** Association of MIND diet and MedDiet with ICAD among people with and without history of MI.

| Model | Model term | Odd Ratio (95%CI) | P Value |
| --- | --- | --- | --- |
| A | History of MI | 20.73 (2.76, 2155.81) | 0.003 |
|  | MIND score | 1.07 (0.95, 1.20) | 0.233 |
|  | MIND x MI | 0.69 (0.53, 0.91) | 0.007 |
| B | History of MI | 49.04 (3.60, 667.29) | 0.003 |
|  | MedDiet score | 1.01 (0.97, 1.05) | 0.757 |
|  | MedDiet x MI | 0.89 (0.81, 0.97) | 0.006 |
| With MI history (n=130) |  |  |  |
| C | MIND score | 0.77 (0.59, 1.01) | 0.053 |
| D | MedDiet score | 0.88 (0.81, 0.97) | 0.010 |
| Without MI history (n=531) |  |  |  |
| E | MIND score | 1.06 (0.94, 1.19) | 0.300 |
| F | MedDiet score | 1.01 (0.96, 1.04) | 0.700 |

death, sex, education, total calories, APOE4 status. Models C-F are the stratified analyses with and without history of MI.

In stratified analyses, the negative association of diet and atherosclerosis was only present in those with history of MI (MIND: OR = 0.77; 95% CI = 0.59, 1.01, p =0.053; Med Diet: OR = 0.88; 95% CI = 0.81, 0.97, p =0.010, Table 3), indicating that healthy diet reduced the risk of atherosclerosis in people with history of MI.

Next, we included an interaction between diet and history of HTN in the model of atherosclerosis as an outcome (Table 4). History of HTN and MedDiet diet interaction term in models of atherosclerosis was significant, indicating that among people with history of HTN the inverse association between MedDiet diet and atherosclerosis is stronger (Table 4). Although in stratified analysis, we did not find any association of either diet score with atherosclerosis (Table 4).

**Table 4:** Association of MIND diet and MedDiet on Atherosclerosis among people with and without history of hypertension (HTN)

| Model | Model term | Odd Ratio (95%CI) | P Value |
| --- | --- | --- | --- |
| A | History of HTN | 7.9 (1.49, 41.61) | 0.015 |
|  | MIND score | 1.16 (0.97, 1.40) | 0.199 |
|  | MIND x HTN | 0.81 (0.65, 1.01) | 0.055 |
| B | History of HTN | 16.4 (1.95, 137.85) | 0.010 |
|  | MedDiet score | 1.04 (0.98, 1.10) | 0.199 |
|  | MedDiet x HTN | 0.93 (0.86, 0.99) | 0.029 |
| With HTN history (n=130) |  |  |  |
| C | MIND score | 0.94 (0.83, 1.07) | 0.404 |
| D | MedDiet score | 0.96 (0.92, 1.01) | 0.126 |
| Without HTN history (n=531) |  |  |  |
| E | MIND score | 1.14 (0.95, 1.37) | 0.160 |
| F | MedDiet score | 1.03 (0.96, 1.09) | 0.400 |

), sex, education (continuous, years), total calories (continuous, kcal/day), and APOE4 status.
<sup>b</sup> Logistic regression models adjusted for age at death (continuous, in years), sex, education (continuous, in years).

Additionally, we ran secondary analyses to examine if the association of MI and HTN with intracranial atherosclerosis varies by low (MIND <7.4 , MedDiet <30) and high diet score (MIND ≥7.4 , MEddiet≥30). The interaction term for MIND diet (MIND (low/high)*HTN: p for interaction=0.49; MIND (low/high)*MI: p for interaction=0.022) and MedDiet (MedDiet (low/high)*HTN: p for interaction=0.055; MedDiet (low/high)*MI: p for interaction=0.024) were assessed. In individuals with low diet scores, MI (MIND diet: OR 1.98, 95% CI 1.18, 3.32; Med diet: OR 2.12, CI 1.25, 3.60) and HTN (MIND diet: OR 1.81, 95% CI 1.15, 2.84; Med diet: OR 2.31, 95% CI 1.41, 3.80) and are associated with higher odds of atherosclerosis. However, in individuals with high diet scores, no association was demonstrated: MI (MIND diet: OR 0.92, CI 0.54, 1.58; Med diet: OR 0.84, CI 0.50, 1.42) or HTN (MIND diet: OR 1.28, CI 0.81, 2.01; Med diet: OR 1.13, CI 0.74, 1.73).

### Sensitivity analysis

We conducted two sensitivity analyses. First, we controlled for other vascular factors with potential to affect associations of interaction between diet and MI and HTN with atherosclerosis. Each of the following terms was added separately to the model with atherosclerosis as the outcome: a history of hypertension, a history of diabetes, a history of stroke, history of smoking, and a history of heart failure. The interaction results between both diets (MIND and MedDiet) and vascular risk factors (MI and HTN) to reduce the risk of atherosclerosis were essentially unchanged (p<0.05; data not shown). Finally, we controlled arteriolosclerosis given that arteriosclerosis is strongly associated with atherosclerosis. We found that the results were also unchanged (p<0.05, data not shown).

## DISCUSSION

In this study of deceased community dwelling older adults, we found history of vascular risk factors, such as MI and HTN, associated with ICAD, but diet was not. Although diet has been found to be beneficial in reducing cardiovascular disease, slowing cognitive decline, and reducing neuroimaging markers of vascular disease, our analysis did not reveal an association between diet and ICAD neuropathology in the overall cohort. However, we found that within individuals with a history of HTN or MI, a healthy diet was associated with lower odds of having ICAD. Additionally, we also found that among those with high diet scores, the association of MI or HTN with ICAD was attenuated. These findings suggest that a healthy diet may benefit in preserving the vascular health of the brain for individuals with pre-existing HTN or history of MI, the conditions associated with ICAD pathology in the brain.

Previous studies have supported the association between diet and ICAD, though human studies of intracranial vascular neuropathology are lacking. In non-human primates, a high-fat and high-fructose (HFF) diet was associated with the intracranial vessel infiltration of lipid-laden foam cells and the appearance of lipid droplet-filled smooth muscle cells, as well as increased vascular reactivity and vasoconstriction to stressors.^24^ In the NOMAS MRI sub study, increased Mediterranean diet scores were inversely related to ICAD, although this trend did not reach significance.^25^ Metabolic syndrome, a common sequela of an unhealthy diet, has been associated with both extra and intracranial atherosclerotic disease on neuroimaging,^26^ with ICAD worsening as the severity of the metabolic syndrome worsens.^27^ A variety of prior studies including ours have found associations between a healthy diet, neuropathology, and functional outcomes, but the focus has been on Alzheimer’s pathology and other structural markers.^28–30^

To the best of our knowledge, this is the first study in humans evaluating the association of dietary patterns during more than 10 years of follow-up with intracranial vascular neuropathology at the time of death. Our analysis examining individuals with low and high diet scores, found that diet may have a protective effect, mitigating the risk of existing vascular conditions on intracranial atherosclerosis, despite a history of diseases known to be strongly associated with the development of atherosclerosis, including HTN and MI. This may suggest that once diagnosed with HTN or MI, diet is of critical importance in protecting brain health via the prevention of intracranial atherosclerosis.

The risk factors and mechanisms for intracranial atherosclerosis are less well understood than for systemic atherosclerosis.^31^ However, two major components of ICAD formation are 1) atherosclerosis due to cholesterol deposition causing inflammation and 2) arterial stiffness resulting from endothelial dysfunction causing sclerosis.^31^ Of the modifiable vascular risk factors, HTN may be the most important factor in the development of ICAD. In Asian and African populations, where the rates of ICAD are highest, the high prevalence of HTN is may partially explain the higher prevalence of ICAD compared to other populations.^7^ Our finding that hypertensive individuals who consume a healthy diet have less cerebral atherosclerosis than those who consume a less healthy diet offers a potential way to mitigate the risk of developing ICAD in high risk populations.

Healthy diets such as MIND and MedDiet, are primarily plant-based, high in fiber, essential nutrients, and antioxidants, and low in sugar and fat. A potential mechanistic link between diet and atherosclerosis is the role of the gut microbiome and inflammation. The intestinal breakdown of dietary proteins, particularly those contained in red meat and egg yolks, creates metabolic products such as carnitine and phosphatidylcholine that cause atherosclerosis and increase the cardiovascular risk in animal models.^32^ The continued breakdown of these metabolites results in the production of trimethylamine N-oxide (TMAO). In a Cleveland Clinic study of individuals undergoing coronary angiograms, those with the highest quartile of plasma TMAO levels had a 2.5-fold increased risk of stroke, myocardial infarction, or vascular death over a three-year follow-up period.^33^ As TMAO and other gut-derived uremic toxins are renally excreted, even mild renal impairment, which is common in individuals with hypertension, can lead to elevated plasma levels.^32^

Another potential mechanism is the contribution of dietary fiber. The fiber content in healthy diets, such as the MIND and Mediterranean diets, is higher than in a Western diet pattern. Fiber acts in multiple ways to improve metabolic health, including the prolongation of stomach emptying, lowering of postprandial blood glucose, and increase of bile excretion, which lowers total and low-density lipoprotein (LDL) cholesterol. Lower total and LDL cholesterol levels have been associated with less atherosclerosis.^34^ Additionally, fiber intake causes stomach distention, which triggers the release of hormones that increase satiation, creating a feedback loop that improves metabolic health. Apo E4 status, best known for its role in Alzheimer’s dementia, has also been associated with a heightened risk of atherosclerosis.^35^ The interplay between genetic risk, diet, vascular risk factors, and neuropathology (both vascular and degenerative) suggests a complex intracranial ecosystem that is just beginning to be fully understood. These hypothetical mechanisms must be further evaluated, particularly regarding intra vs. extracranial atherosclerosis and potential variations related to the blood–brain barrier and other mediating factors.

The strengths of this study include a large number of autopsies, standardized neuropathological measures, use of a validated food frequency questionnaire administered annually, and structured clinical assessments from study enrollment until death. These measures reduce the selection bias resulting from loss to follow-up and the potential for measurement error. The limitations of this work include its observational study design, which precludes our ability to claim causal relationships. The participants were mostly White, non-Hispanic older individuals, so caution should be exercised in generalizing to more diverse populations or to younger adults. Future studies should explore potential mechanisms, particularly the role of inflammation and biomarkers, and the interplay between vascular neuropathology and other brain pathologies.

## Conclusion

In older adults with a history of hypertension or myocardial infarction, a healthy dietary pattern is associated with less severe intracranial large-vessel atherosclerosis. These data suggest that diet may be effective in maintaining vascular brain health in persons with a history of vascular risk factors. In-vivo studies of dietary habits and brain health are needed, specifically in those at high vascular risk.^36^

## ACKNOWLEDGMENTS

We thank the participants of the Rush Memory and Aging Project and key staff members; Traci Colvin, MPH, for coordination of the clinical data collection; Karen Skish, MS, for coordination of the pathologic data collection; John Gibbons, MS, and Greg Klein, MS, for data management. The study was funded by NIH (R01AG17917, P30AG10161, R01AG15815, R01AG34374) and the Illinois Department of Public Health. The funding organizations had no role in the design or conduct of the study; the collection, management, analysis, or interpretation of the data; or the writing of the report or the decision to submit it for publication.

## DATA AVAILABILITY STATEMENT

All data in these analyses (and descriptions of the studies and variables) can be requested through the Rush Alzheimer’s Disease Center Research Resource Sharing Hub at www.radc.rush.edu.

